# Blood Culture Utilization Optimization: Data-Driven Approaches to Guide Stewardship During Critical Supply Shortages

**DOI:** 10.1101/2024.10.02.24314807

**Authors:** Fatemeh Amrollahi, Nicholas P. Marshall, Amy Chang, Stanley Deresinski, Niaz Banaei, Stephen Ma, Jonathan H. Chen

**Affiliations:** Biomedical Informatics, Stanford University Palo Alto, CA 94304, USA; Division of Pediatric Infectious Diseases, Stanford University Palo Alto, CA 94304, USA; Stanford University Palo Alto, CA 94304, USA; Clinical Microbiology Laboratory, Stanford University Palo Alto, CA 94304, USA; Division of Hospital Medicine, Stanford University School of Medicine, Palo Alto, CA 94304, USA

## Abstract

Amidst the global blood culture bottle shortage, there is an urgent need to reduce unnecessary blood cultures. This study developed BSICalc and BSIScore, data-driven models using electronic health record (EHR) data, to guide blood culture ordering in the emergency department at the point-of-care, enhancing stewardship efforts and supporting clinical decision-making.

## Introduction

There is a global shortage of blood culture bottles, creating an urgent need to optimize their use.^1,2^ Blood cultures are the gold standard for diagnosing blood stream infections (BSI), yet fewer than 10% yield positive results, with up to 60% potentially ordered without clear clinical indications.^3,4^ Unnecessary blood cultures carry significant downsides, including the risk of contamination, which can lead to unnecessary hospitalization, antibiotic use with associated side effects, and additional phlebotomy and laboratory testing, further straining an already burdened healthcare system.^5,6^ This shortage has heightened the urgency to reduce unnecessary blood cultures, ensuring that limited supplies are allocated to patients with the most clinically appropriate indications.^1,2^ Expert organizations have issued guidance to help prioritize high-yield blood cultures,^4,6^ but applying these recommendations remains challenging, particularly in high-pressure environments like emergency departments (EDs), where most blood cultures are ordered. Previous study efforts to guide blood culture collection using decision rules based on clinical factors have shown promise but were often limited by small sample sizes and variable clinical interpretation.^5,6^ This study aims to develop and validate data-driven decision rules using a large dataset from an electronic health record (EHR) system across academic and community hospital settings, facilitating automated and integrated clinical decision support.

## Methods

We conducted a retrospective analysis of EHR data from 135,483 ED blood culture orders, with the primary outcome being a positive result. The study included patients aged 18 years or older who had blood culture sampling during their ED visits, provided they had no positive blood cultures within the preceding 14 days. Cultures that were marked with errors, discontinued, or canceled were excluded from the analysis. The primary outcome of interest was a positive blood culture result, with exclusions for contaminants identified in multiple bottles by laboratory protocols and those with coagulase-negative staphylococci or gram-positive rods. For this study, to ensure broad applicability across various healthcare systems, we utilized primary laboratory and clinical variables commonly measured in care settings. The data was split chronologically into training (Culture orders taken between 2015-2022), development (culture orders taken between 2022-2023), and evaluation sets (culture orders taken 2023 onwards), allowing the most recent data to be used for evaluation and providing a more accurate representation of system performance in real-world settings. We developed BSICalc, a logistic regression model (Equation 1) to predict blood culture outcomes, and derived BSIScore, a point system based on lab results and vital signs within 24 hours of the order described below. Each feature was dichotomized using median values. A cumulative score ≥4, with a sensitivity of 0.95, indicated a high likelihood of a necessary blood culture order leading to a positive result.

- Maximum Heart Rate >100 BPM (+2 points)
- Maximum Temperature >100.4°F (+3 points)
- Minimum Systolic Blood Pressure <113 mmHg (+2 points)
- Maximum White Blood Count >9.9 × 10^9^/L (+2 points)
- Minimum Sodium <136 mEq/L (+1 point)
- Minimum HCO3 <24 mEq/L (+1 point)
- Minimum Platelet <222 × 10^9^/L (+2 points)
- Maximum Creatinine >0.96 mg/dL (+1 point)
- Maximum Lactate >1.43 mmol/L (+1 point)
- Age >65 (+2 points)

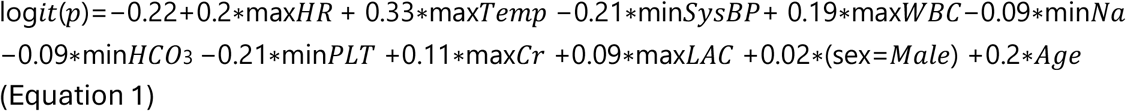

## Results

We benchmarked BSICalc and BSIScore against established models by comparing negative predictive value (NPV), specificity, and positive predictive value (PPV) at a fixed sensitivity of 0.95. Table 1 provides a detailed comparison of these performance metrics.

**Table 1:** This table compares the performance of BSICalc and BSIScore against various SIRS-based criteria and the numerical components of the Shapiro method. The evaluation metrics include sensitivity, specificity, negative predictive value (NPV), and positive predictive value (PPV). BSICalc, with an AUC of 70%, demonstrated high sensitivity (95%) and higher specificity (17%) compared to SIRS criteria, making it a reliable tool for ruling out bacteremia when the test result is negative.

|  | Sensitivity | Specificity | PPV | NPV |
| --- | --- | --- | --- | --- |
| BSICalc | 95% | 17% | 3% | 99% |
| BSIScore (4+ point) | 95% | 17% | 3% | 99% |
| BSIScore (3+ point) | 97% | 9% | 3% | 99% |
| BSIScore (2+ point) | 99% | 3% | 3% | 99% |
| SIRS (2+ criteria) | 78% | 39% | 4% | 98% |
| SIRS (2+ or SBP <90 or Lactate >2.0) | 84% | 32% | 4% | 99% |
| SIRS (1+ criteria) | 95% | 13% | 3% | 99% |
| SIRS 1+ or SBP <90 or Lactate >2.0 | 95% | 12% | 3% | 99% |
| Shapiro (2+ criteria) | 55% | 69% | 5% | 98% |
| Shapiro (1+ criteria) | 87% | 27% | 3% | 99% |

We further evaluated the performance of BSIScore by measuring specificity at various sensitivity levels (see Table 1). This approach allows clinicians to adjust the decision threshold according to the desired sensitivity in their clinical setting, enabling them to select a decision probability that best suits their needs.

## Discussion

Blood culture stewardship is increasingly critical, particularly during the current supply chain crisis. Our study introduces a data-driven approach that enhances clinician judgment by identifying patients that are least likely to have positive blood culture and benefit from blood cultures, thereby reducing unnecessary testing.

We derived a predictive model using readily available EHR data, including vital signs and lab results, to assess variations in SIRS criteria for predicting blood culture positivity. This model was then converted into a user-friendly scoring system, enabling clinicians to apply it at the point of care. Unlike previous decision rules requiring more manual input, our approach identifies patients at low-risk for blood stream infections and allows clinicians to adjust sensitivity based on their clinical setting.

The model’s large sample size enhances its robustness, though the single-site design may limit generalizability. While there is a risk of overfitting, the model’s simplicity and reliance on common EHR data make it practical for real-world use, either as a point-of-care tool or integrated into EHR systems for automated decision support.

In conclusion, data-driven decision rules provide a more precise and reliable method for guiding blood culture stewardship and safety measures, offering a more targeted alternative to the broad rationing strategies currently necessitated by the supply chain crisis.

## Data Availability

Research data available via the Stanford medicine research data repository (https://starr.stanford.edu/).

## Author Contributions

Dr. Amrollahi and Dr. Marshall contributed equally as co–first authors. Dr. Chen had full access to all of the data in the study. All authors were involved in the original conception and design of the work. Dr. Amrollahi developed and performed the analysis and conducted the experiments. Dr. Marshall, Dr. Ma, and Dr. Chen reviewed the experiments and contributed to the interpretation of the results. Dr. Chen, Dr. Chang, Dr. Deresinski, and Dr. Banaei provided clinical expertise in the design and interpretation of the results. Dr. Marshall, Dr. Amrollahi, and Dr. Chen wrote and edited the final manuscript.

## Additional Contributions

We extend our gratitude to Dr. Fateme Nateghi Herdasht, PhD, and Dr. Manoj Maddali, MD, Dr. Sanjat Kanjilal (MD,MPH), Dr. Richard Medford, MD, Dr. Mary Kane Goldstein MD, Dr. Steve Asch (MD, MPH), Dr. Tho Pham, and Dr. Jorge Salinas for insightful discussions and assistance in conceptualizing the problem.

